# Exploring two-way text messages for post-discharge follow-up and quality improvement in rural Uganda

**DOI:** 10.1101/2025.04.02.25325125

**Authors:** Charly Huxford, Bella Hwang, Dustin Dunsmuir, Yashodani Pillay, Fredson Tusingwire, Florence Oyella Otim, Beatrice Akello, Aine Ivan Aye Ishebukara, Stefanie K Novakowski, Bernard Opar Toliva, Nathan Kenya-Mugisha, Abner Tagoola, Matthew O Wiens, Niranjan Kissoon, J Mark Ansermino

## Abstract

**Introduction:** Automated messaging through text (SMS) and instant messaging services (IMS) are low-cost solutions for patient follow-up in resource-constrained contexts. This study aims to evaluate a quality improvement (QI) initiative to improve caregiver response rates to an automated messaging system facilitating follow-up after hospital discharge of children in rural Uganda.

**Methods:** This initiative was implemented at Gulu Regional Referral Hospital in Northern Uganda from June 2022 to June 2024. Caregivers of children who were triaged through the Smart Triage digital platform were offered an automated follow-up program as part of routine care during this period. SMS and IMS (WhatsApp) messages prompting caregivers to report if their child had “improved” or “not improved” were sent seven days post-discharge. Non-responders and "not improved" cases were escalated to a phone call from a health worker. From April 2023 to June 2024, a QI initiative refined the messaging system to improve response rates. Data on message delivery, response rates, improvement strategies, and health outcomes were analyzed.

**Results:** Of 6826 participants, 6469 (95%) messages were successfully delivered. Response rates improved from 20% to 40%. In total, 1856 caregivers responded to the messages. Among the responses, 1244 (67%) of caregivers reported improvement and 612 (33%) reported no improvement. Follow-up phone calls for those “not improved” revealed 58 (9%) sought care, 12 (2%) were readmitted, and no deaths occurred. For non-responders, 206 (5%) sought care, 33 (0.7%) were readmitted, and 3 (0.07%) deaths occurred.

**Discussion:** Automated two-way text messages for post-discharge pediatric follow-up in Uganda yielded high delivery but moderate response rates. Iterative QI efforts increased response rates, highlighting the importance of tailored communication strategies. Automated messages can facilitate timely intervention for high-risk children and enable efficient collection of health outcomes offering a viable alternative to in-person follow-up in resource-poor settings.

## Introduction

Post-discharge mortality often exceeds in-hospital rates in low-resource settings [1]. The causes of post-discharge deaths are multi-factorial and include poverty, inability to access timely care, non-resilient health systems, and poor follow-up after discharge. Follow-up of all children post-discharge may not be feasible or even necessary; however, it is certainly a need among those at high risk for post-discharge death. In many cases in-person follow-up is resource intensive and impractical and hence largely ignored or poorly conducted, leaving patients and families with little support during the vulnerable post-discharge period. Our group developed, validated, implemented, and evaluated a digital triage platform called Smart Triage [2–6] to prioritize initial emergencies at facility level. Leveraging data from this platform, we sought to explore ways of improving communication and follow-up after discharge using Short Message Service (SMS) and Instant Messaging Service (IMS) communication.

Previous studies have demonstrated the usability and acceptability of SMS-based interventions, and their potential to enhance treatment adherence and healthcare communication in low-resource settings; however, their effectiveness for structured post-discharge follow-up in pediatric populations remains largely unexamined [8,9]. Mobile phone penetration has risen dramatically in sub-Saharan Africa, with an estimated 49% of the population owning a smartphone and 82% having access to basic mobile phones [7]. SMS and WhatsApp were selected for facilitating follow-up due to their widespread availability and frequent use in daily life. In Uganda, texting is the most common activity among mobile phone users [18]. These messaging platforms offer low-cost, scalable solutions for engaging caregivers, alleviating the burden on overextended health workers, and collecting actionable data in real time. However, limited access to technology among the lowest income households, varying literacy levels, and inconsistent network coverage remain barriers to using text messaging for post-discharge follow-up in resource-poor settings. Furthermore, cultural and contextual factors may influence caregiver engagement and response rates, necessitating a tailored approach to ensure successful implementation [10].

This study aimed to evaluate the implementation of a quality improvement (QI) intervention to increase response rates to automated two-way text message follow-ups for children discharged from Gulu Regional Referral Hospital (GRRH) in Northern Uganda. Additionally, the study explored the potential of using these text messages to support real-world outcome monitoring of a pediatric clinical prediction model and improve communication between caregivers and health workers.

## Methods

### Study design and setting

The study was conducted in the outpatient department (OPD) of GRRH over a two-year period, from June 23, 2022, to June 30, 2024. GRRH is a public hospital funded by the Uganda Ministry of Health and its OPD serves as a primary healthcare provider for over 23,000 pediatric and adult patients annually, offering free services to residents of five districts in Northern Uganda: Amuru, Gulu, Kitgum, Lamwo, and Pader.

The follow-up intervention augments the Smart Triage platform, developed over the past decade, which aims to collaboratively develop and implement clinically validated tools to allocate resources more efficiently in low-resource contexts and improve outcomes for children with an acute illness upon arrival to outpatient (emergency) departments. The Smart Triage platform uses a data-driven pediatric triage risk prediction model and treatment tracking system for children under 18 years old [2–6] and was implemented as a part of routine care. Initially, all caregivers were enrolled during triage for telephone follow-up with a study nurse seven days after leaving the hospital. The follow-up phone calls were supplemented with the automated two-way text messaging system in June 2022.

### The Intervention

#### Automated follow-ups

The automated two-way follow-up aimed to assess the health status of children seven days after triage and their subsequent discharge directly from outpatient (emergency) care or seven days after discharge from inpatient care. During triage, caregivers were asked if they would like to receive an automated follow-up. If consent was provided, caregivers selected their preferred communication channel—either SMS or WhatsApp—and their preferred language. SMS messages were available in both English and Acholi, while WhatsApp messages were offered in English only. The follow-up message asked caregivers for a simple binary response at no cost to them: *"A health worker from Gulu Hospital is following up with you to see if your child [child’s name] has improved. Please reply with ’1’ if your child has improved or ’2’ if your child has not improved. This number is toll-free. Send STOP or 196 to unsubscribe."*

The automated follow-up was used to immediately identify children who had not improved. Caregivers who responded that their child had not improved were flagged for further follow-up by a study nurse, who contacted them by phone as soon as possible. A reminder message was automatically sent if caregivers did not respond within three days of receiving the initial message. Non-responders (those who did not reply within seven days), caregivers who unsubscribed (those who responded with STOP or the number 196), and caregivers who sent an invalid response (a text-based response other than 1 or 2) were also contacted by phone to assess the child’s status. Those who responded that their child had improved were sent an automated acknowledgment message, with no further follow-up. Caregivers who were unreachable by phone call after multiple attempts were considered lost to follow-up.

#### Automated follow-up system technology

Data included follow-up dates, phone numbers, responses to the messages, and responses to the phone call questions and were stored in a Research Electronic Data Capture (REDCap) database. REDCap is a secure and encrypted, web-based application designed for data collection and management in research studies and quality improvement projects [13]. REDCap required 2-factor authentication to access to maintain the security of caregiver and patient contact information. The automated follow-up system used Africa’s Talking (https://africastalking.com) for SMS and Twilio (https://www.twilio.com/en-us) for WhatsApp messaging. Seven days after discharge, messages were sent through the caregiver’s preferred channel—SMS or WhatsApp—using a scheduled server process that triggered message delivery daily at 4:00 PM (originally 9:00 AM) East Africa Time. Caregiver responses to messages were processed by a dedicated callback URL which logged and updated REDCap. Automatic logging and daily monitoring by the study team ensured messages were sent and delivered. Responses were automatically tracked within REDCap. Issues with message delivery, such as failed WhatsApp messages, were also recorded automatically on REDCap. Failed WhatsApp messages were automatically resent as an SMS message.

#### SMS

Africa’s Talking was used for SMS follow-ups through a dedicated short code (4-digit phone number available in Uganda). The SMS delivery server application used the Africa’s Talking Python library (version 1.2.7) and caregiver responses were managed through callbacks to a Flask-based Application Programming Interface (API) that then updated the REDCap database.

#### IMS

Caregivers who selected WhatsApp as their preferred communication method received automated messages via Twilio. All message text was required to be pre-approved by Twilio and so could not be sent in the local language. The local language, Acholi, has a relatively small number of native speakers and was not an included language on the Twilio platform. The Twilio Python server-side software development kit (SDK) was used to initiate conversations based on REDCap data and then a flow created in Twilio Studio controlled the conversation logic and handled responses, including updating to the REDCap research database.

#### QI to increase the message response rate

The baseline response rate was established between June 2022 to March 2023, before the implementation of any QI changes. From April 2023, multiple iterative strategies were implemented on a monthly basis to improve response rates and refine the messaging process (Figure 1). Response rates were monitored over time to assess the impact of each intervention. The study’s systematic introduction of response strategies allowed for adjustments based on feedback and evolving needs, aiming to improve follow-up response rates.

**Figure 1.**
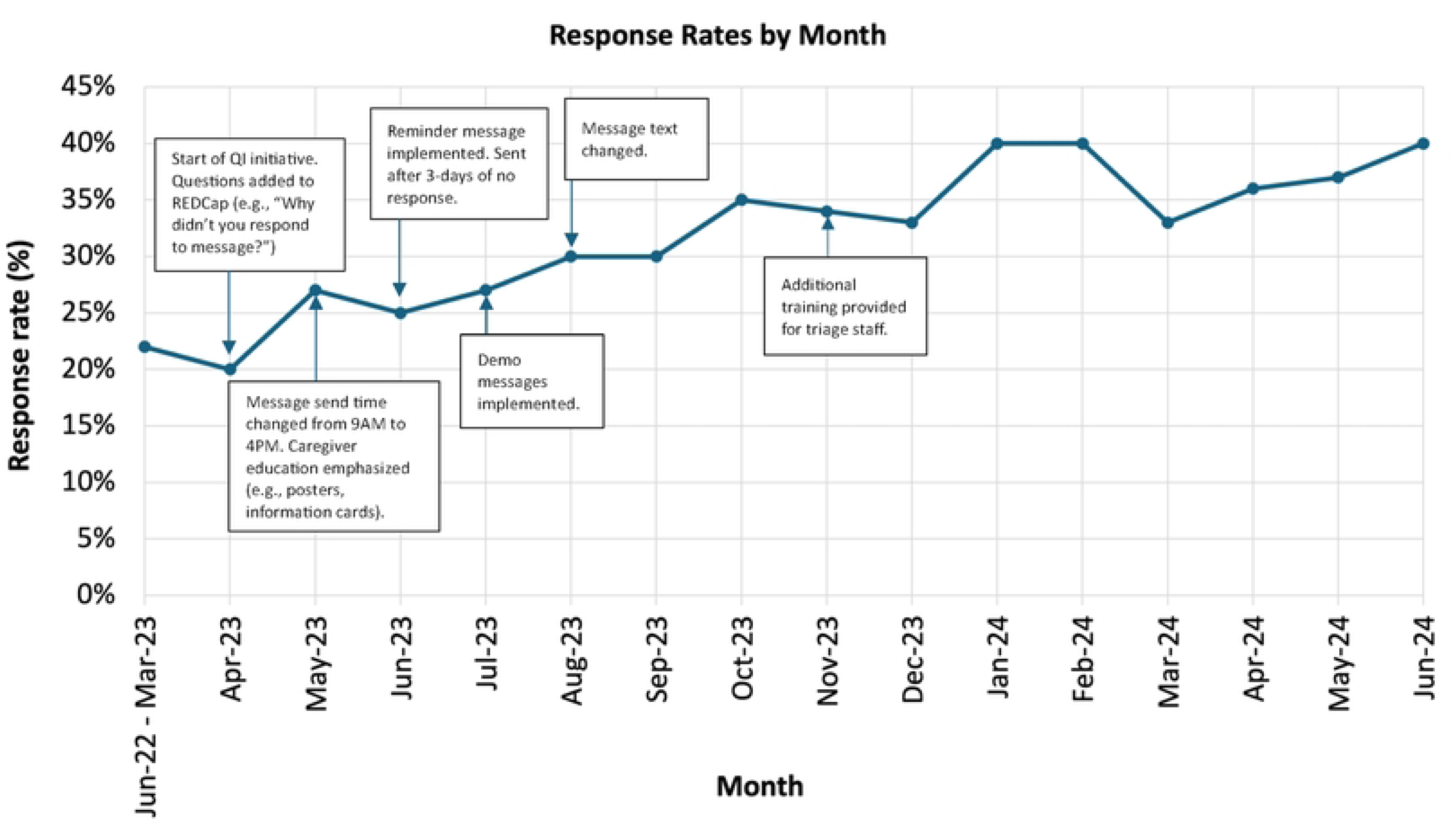
Response rate change by month

### Data collection

Responses to the automated messages were automatically recorded in REDCap in real-time, ensuring accurate and timely data entry. If a caregiver responded with "1" (improved) the case was marked as complete and sent an acknowledgment. Responses of "2" (not improved) and non-responses were tracked in a REDCap report, which was monitored daily. The study nurses attempted to call all caregivers in this report. During the phone call, caregivers were asked a standardized set of questions about their child’s health and the study nurses manually entered the responses into REDCap immediately following the interaction.

### Outcome measures

#### Delivery rate

The success of the automated follow-up message is described by the delivery and response rates as percentages. Message send attempts may fail to be delivered due to a telecom outage, the number being not in service, or, for WhatsApp, the number not being connected to an account. The delivery rate was the number of messages delivered out of the number of messages sent.

#### Response rate

The response rate was the number of messages replied to out of the number of messages delivered. A response was defined as the caregiver responding that their child had improved or not improved. During the follow-up phone calls, caregivers who did not respond to the text messages were asked for their reasons, which were broadly categorized into technology-related issues and caregiver-related factors. Reasons for non-response were unavailable in cases where caregivers were unreachable, reported having replied, or the reason was unspecified.

#### Health outcomes

Health outcomes were collected with a standardized set of questions asked by the study nurse during the phone call follow-up. The questions about their child’s condition included whether they had sought additional medical care without readmission, if the child had been readmitted, and if the child had died. The collected data were analyzed to determine the proportion of children experiencing each outcome.

### Ethics

Ethics approval for this study was obtained from the institutional review boards at Makerere University School of Public Health in Uganda (SPH-2021-41) and the Uganda National Council for Science and Technology (HS1745ES). As this is a quality improvement project, patient consent was waived. Ethics approval for this study was not required by the University of British Columbia’s (UBC) Research Ethics Board. UBC adheres to the Canadian government’s <u>Tri-Council Policy 2 (TCPS2) Statement</u> which states that quality assurance and quality improvement (QA/QI) studies, program evaluation activities, and performance reviews, or testing within normal educational requirements, when used exclusively for assessment, management or improvement purposes, do not constitute research under the TCPS 2 and do not fall under the scope of REB review. See section 4.4.1 of the <u>UBC Clinical Research Ethics</u> <u>General Guidance Notes.</u> No data was collected in Canada. This QI project was reported according to the Standards for Quality Improvement Reporting Excellence (SQUIRE) 2.0 guidelines [20].

## Results

### Delivery rate

There were 6826 caregivers enrolled in the automated follow-up system (Figure 2). In total, 5715 (84%) caregivers chose SMS as their preferred channel and 1111 (16%) chose WhatsApp. The overall delivery rate was 95%. Of the 357 (5%) undelivered messages, most (98%) were WhatsApp messages, while 2% were SMS messages.

**Figure 2.**
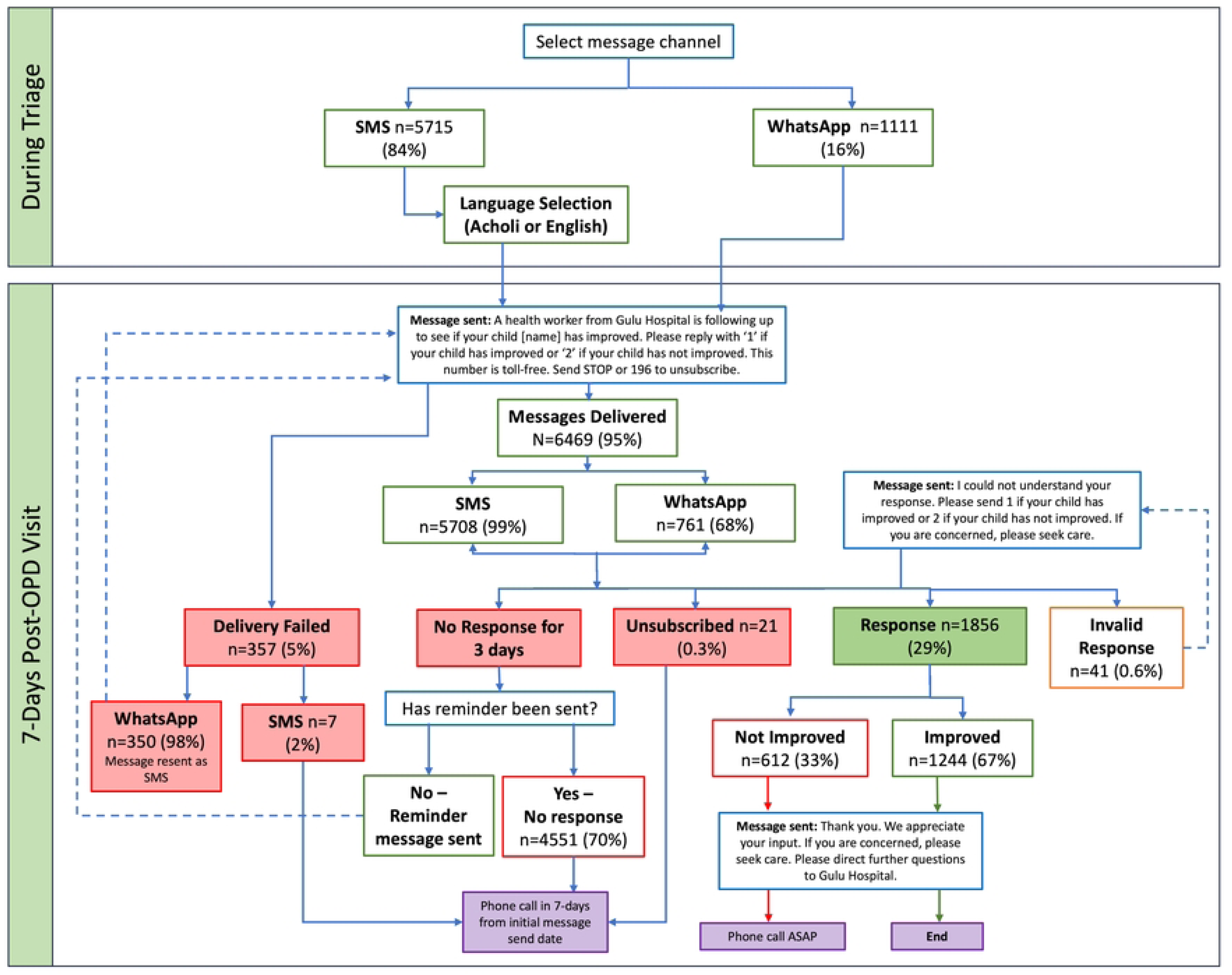
**Messages sent and received**

### Response rate

Between June 2022 and March 2023, before the implementation of any QI changes, the response rate was 22% (507/2292). In April 2023, at the start of the QI intervention, the response rate was 20% (79/402) which steadily increased to 40% (107/265) by June 2024 after a series of iterative changes. Between June 2022 and June 2024, 1856 responses were received. Out of those, 1244 (67%) caregivers responded that their child had improved, and 612 (33%) responded that their child had not improved.

The QI changes implemented were informed by the feedback from caregivers (Table 1). The data gathered through follow-up calls with non-responders identified key barriers such as message delivery times, confusion about the sender, and lack of understanding of how to respond. For example, follow-up messages were initially sent at 9:00 AM. After receiving feedback that caregivers were often busy in the mornings, the message time was shifted to 4:00 PM to better align with their availability. Another strategy was to add a reminder message if no response was received three days after the initial message, to prompt caregivers who may have forgotten or missed the first message. In addition, to ensure caregivers understood how to respond, a demo message was sent to the caregiver’s phone at the time of their in-person triage. A health worker demonstrated and explained the follow-up message to caregivers. Additionally, caregiver education was enhanced using posters displayed in the OPD and information cards distributed during triage. The posters and cards provided additional information regarding the automated follow-up system, including the definitions of the response options and understanding the response process.

**Table 1.** Reasons for not responding n=1524.

| Reason participant did not respond | n (%) |
| --- | --- |
| Technical issue (e.g. broken phone) | 588 (39) |
| Did not know how to reply to the message | 267 (18) |
| Busy | 230 (15) |
| Forgot to respond | 223 (15) |
| Worried about the cost to respond | 128 (8) |
| Did not know who was sending the message | 26 (2) |
| Not interested | 15 (1) |
| Could not be reached at number/is no longer available (caregiver provided number of relative, neighbor, friend, etc.) | 6 (0.4) |
| Wrong number (caregiver gave wrong number or recipient does not know the caregiver) | 5 (0.3) |
| Other<sup>a</sup> | 36 (2) |

<sup>a</sup>Other includes reasons such as not having data, phone was off, didn't take it seriously, lost phone, didn't check messages

### Health outcomes

A total of 5582 caregivers were scheduled for follow-up phone calls by the study nurse. Those scheduled included 4551 caregivers (82%) who did not respond to the automated message, 612 (11%) who replied that their child had not improved, 41 (0.7%) who sent an invalid response, 21 (0.4%) who unsubscribed, and 357 (6%) for whom the message delivery failed (Figure 2). As part of the protocol, caregivers who unsubscribed from the messages were still followed up with by phone call. A total of 5619 caregivers were actually reached by phone, including 177 caregivers (3%) who received a follow-up phone call even though they responded that their child had improved (calls were not required for this group). This occurred due to either delayed caregiver responses, which arrived after the follow-up call had already been made, or errors by the study nurse. In total, 140 caregivers (3%) were lost to follow-up.

Among the 4551 non-responders, 206 caregivers (5%) reported seeking care, 33 (0.7%) stated their child had been readmitted, and 3 children (0.07%) had died by the time of follow-up (Table 2). Among the 612 caregivers who reported that their child had not improved, 58 (9%) indicated they had sought some care that did not lead to an improvement, 12 (2%) reported that their child had been readmitted, and none had died by the time of the phone call. The most common reasons for readmission were malaria (5 cases, 41%) and upper respiratory tract infections (URTI; 2 cases, 17%).

**Table 2.**
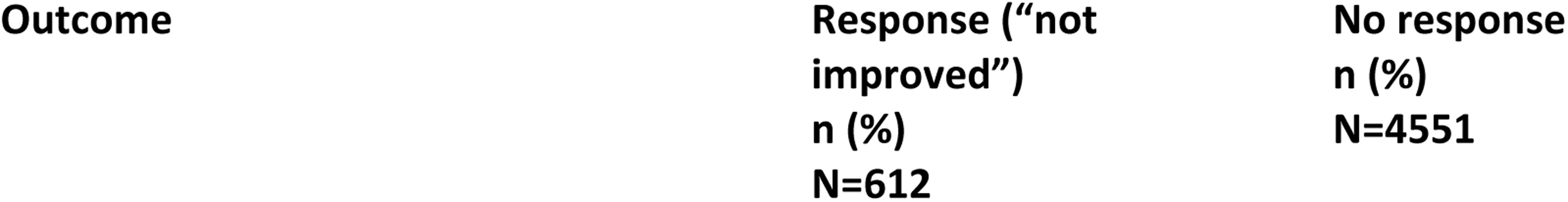
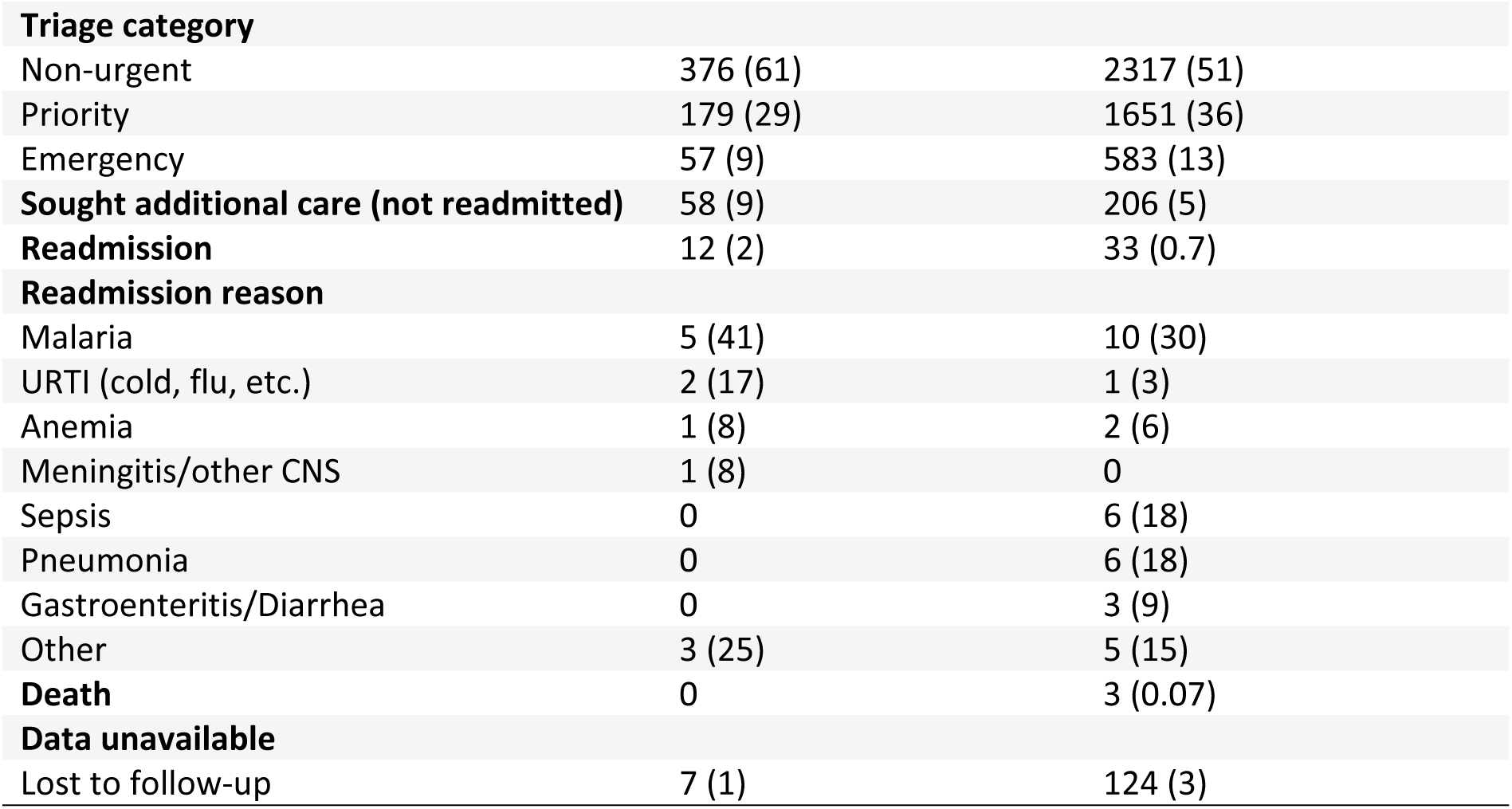
Health outcomes collected during follow-up phone calls.

## Discussion

The automated text messaging system achieved a high delivery rate (95%), with the majority of undelivered messages (98%) sent via WhatsApp. Caregiver response rates increased to 40% from 20% at the start of the initiative. Iterative strategies, such as optimized message timing, reminder messages, and enhanced caregiver education played a critical role in these improvements. Strategies to address non-response to follow-up messages should consider context, available resources, and risk assessment. Our findings suggest that non-responders had fewer adverse outcomes than those who responded, indicating that failure to respond may often reflect a lack of caregiver concern due to perceived improvement in the child’s condition. A selective approach to following up with non-responders that prioritizes those identified as high-risk during hospital triage or focuses efforts where there is a higher likelihood of adverse outcomes may be reasonable, although further research would be needed to assess such an approach. Such a strategy would balance resource constraints with the need for effective care, allowing for regionally and contextually appropriate interventions tailored to what is affordable and achievable within the local healthcare infrastructure.

We used SMS and IMS as simple and effective solutions for post-discharge follow-up in children. Each solution has advantages that will evolve with advances in technology availability. SMS is more widely available and is supported by feature phones but requires a relay service. IMS requires a smartphone and data plan, but it is more reliable as it does not involve the local telecom and can be delivered at a lower cost and at scale. By combining an automated two-way messaging system with an iterative QI approach, we systematically addressed barriers to caregiver engagement and achieved sustained improvements in response rates over the study period. While the peak response rate of 40% may appear modest, it is important to note that this encompassed all children who were triaged, rather than focusing solely on those at higher risk. A 100% response rate may be unrealistic when enrolling all discharged children due to systemic barriers such as limited mobile network coverage and caregiver concerns about the cost of responding. The messaging system can be combined with a risk prediction model to identify non-responders most in need of a follow-up call by a human. This approach would help ensure that the low response rate is less of a concern by distinguishing between those who are likely well and those who may require care, including caregivers who should have sought medical care for their children but did not.

### Comparison with other studies

SMS-based interventions are effective in healthcare settings in low-and middle-income countries (LMICs) and are reported to improve childhood immunization coverage and timeliness [10], increase attendance at maternal and neonatal healthcare visits [11], and support treatment adherence in the management of hypertension and HIV [12, 8]. However, many studies using text messages have primarily focused on sending reminders or behavior change interventions aimed at improving specific outcomes, such as appointment attendance and medication adherence, and mostly in non-urgent conditions where there is time for deliberation [14].

Strategies to improve response rates for text-based interventions include personalizing messages, such as incorporating the patient’s name or tailoring content to specific health conditions [15, 16]. Sending messages at times aligned with recipients’ routines or preferences also improves response rates [15, 16]. In our study, strategies such as personalizing messages to include the child’s name and adjusting the timing of message delivery were implemented, leading to an overall increase in response rates.

The potential for text-based interventions to improve post-discharge outcomes is highlighted by a study conducted in rural Kenya. The study evaluated the use of SMS for following up with caregivers of children after discharge. The findings revealed improvements in caregiver knowledge, increased efficacy in childcare, strengthened relationships with healthcare staff, and reduced hospital readmission rates [17].

### Strengths and limitations

The strength of our study is the iterative QI approach which tailored the system for the environment in which it was routinely used. We drew on successful strategies from previous studies, including the WelTel Kenya1 study [8], which demonstrated the effectiveness of using simple messages such as “How are you?” to encourage responses and enable self-reporting of problems. Building on this approach, our system utilized two-way text messaging to confirm whether caregivers received and engaged with the messages and to capture valuable feedback on the health of children once they returned to their community. This feedback may allow health workers to respond quickly when caregivers report a lack of improvement in their child’s condition, potentially preventing adverse outcomes.

There is a critical need for community post-discharge outcome data and follow-up care for high-risk children, as well as a more efficient approach to reduce the burden on health workers and healthcare resources. Before the introduction of the automated follow-up system, health workers attempted to call every caregiver of a discharged child—a highly resource-intensive process, with each call taking at least five minutes. A more targeted approach, in which follow-up calls focus only on caregivers who do not respond to messages or report that their child has not improved, can streamline efforts and make follow-up more feasible. While individual phone calls remain time-intensive, prioritizing high-risk cases using data-driven strategies can reduce overall resource demands compared to traditional methods. Additionally, the system ensures timely data collection, which is essential for driving QI initiatives and refining clinical risk prediction models. The growing response rates, along with the simplicity and accessibility of text messaging, make it a viable alternative to traditional follow-up methods—such as return visits—particularly in settings with limited infrastructure and long travel distances. Beyond facilitating follow-up, the system captures valuable community-level outcomes, such as readmissions and post-discharge morbidity, providing crucial data for clinical and public health planning. Furthermore, the collected data can help identify key trends, including seasonal variations in disease prevalence and shifts in care-seeking behaviors, which can inform targeted public health interventions.

Limitations are related to the technology and barriers faced by caregivers. Undelivered messages due to rurality and infrastructure (e.g., caregivers lacking active accounts or compatible devices, inconsistent mobile network coverage, and occasionally disrupted message delivery) were significant, given the total number of messages sent. Caregiver-related barriers included difficulty understanding the messages, competing priorities, unawareness of the sender or purpose of the message, and concerns about cost (although they were assured that this is not an issue). Incorrect or outdated phone numbers and broken screens and keypads were also problems. These findings highlight the importance of addressing behavioural and technological obstacles to improve response rates.

The costs associated with this system are relatively high in the Ugandan context, with fixed monthly and annual payments to Africa’s Talking and the Ugandan Communications Commission amounting to USD $3,776.00 per year. These costs are fees paid to the mobile network operators and for the use of a dedicated short code. Although the cost per individual message was low— USD $0.016 for outgoing WhatsApp and USD $0.0018 for incoming and outgoing SMS—these expenses can be costly at scale and accumulate significantly with high patient volumes. Twilio charges a small per-message fee for each outgoing WhatsApp message, which decreases as the volume of messages increases. However, the system could be cost-effective if it is shared across multiple facilities, distributing the fixed costs more efficiently.

Mobile applications like WhatsApp offer a more cost-effective solution for communication compared to SMS. However, in our study, most caregivers preferred SMS as their communication channel. This preference may be attributed to the study’s setting in a relatively poor region of Uganda, where access to smartphones is likely limited for many caregivers. In the Acholi region which encompasses Gulu district, 68% of households owned a mobile phone compared to 93% in higher income areas like Kampala [19]. Despite these costs, the system has the potential to enable efficient and cost-effective care if it reduces hospitalizations by improving follow-up. Leveraging predictive models [3, 4], to identify and target follow-ups for the most vulnerable children could maximize the impact of an automated follow-up system while minimizing unnecessary expenditures and could be a valuable investment in resource-constrained settings.

## Conclusion

Automated two-way text messaging systems for post-discharge follow-up in children using SMS and IMS to connect healthcare workers with caregivers is feasible. Response rates improved as a result of QI initiatives that included refining the intervention and addressing real-world barriers and caregiver needs. This study highlights the potential of integrating digital health technologies in resource-limited settings where traditional follow-up methods may be impractical or overly resource-intensive. The potential to scale this approach offers opportunities for broader applications in healthcare, supporting a shift toward digital health innovations in resource-limited contexts. This approach could be further adapted and scaled to include additional functionalities, such as interactive chat features and expanded language options, improving accessibility. The system may be tailored to minimize time-intensive phone calls by prioritizing children initially triaged as emergencies or caregivers who report that their child has not improved. Continued efforts to optimize response rates, address technological limitations, and improve accessibility are essential to maximizing the potential of digital health interventions for enhancing follow-up care in resource-constrained settings. Work is ongoing with the Uganda Ministry of Health to integrate this program within their system.

## Data Availability

Study materials (dataset, data dictionary, and metadata) are publicly available through the Pediatric Sepsis Data CoLaboratory’s (Sepsis CoLab) Dataverse: https://doi.org/10.5683/SP3/XSLNKY. Due to the sensitive nature of clinical data and the potential risk for re-identification of research participants, the de-identified dataset is available through moderated access. Access to this data will be granted on a case-by-case basis following approval from the authors and the Data Governance Committees.

## Acknowledgments

We are grateful for the efforts of facility staff and leadership at Gulu Regional Referral Hospital. Further, we would like to thank members of the Smart Triage research team based at Walimu, Uganda and the Institute for Global Health, Canada. This includes but is not limited to, Savio Mwaka, Clare Komugisha, Bamwesigye Emmanuel, Isaac Omara, Nainigino Marion Naunah, Reagan Obalim, Victor Kissa, Peter Lewis, Jessica Rigg, Katija Pallot, Parnian Hosseini, Alishah Mawji and Edmond C K Li. We are also immensely grateful for the support from the BC Children’s Hospital Foundation and Mining4Life.

